# Pooled testing for SARS-CoV-2 surveillance in schools: real-world evaluation of transmission control, testing resources, and educational disruption

**DOI:** 10.64898/2026.06.03.26354821

**Authors:** Elisabetta Colosi, Lucille Calmon, Montserrat Fässli, Katrin N. Koch, Julia A. Bielicki, Vittoria Colizza

**Affiliations:** Bocconi University, Dondena Research Centre for Social Dynamics and Public Policy, Milan, Italy; Sorbonne Université, INSERM, Pierre Louis Institute of Epidemiology and Public Health, Paris, France; ISI Foundation, Turin, Italy; Paediatric Research Centre, University Children’s Hospital of both Basel (UKBB), Basel, Switzerland; Cantonal Office of Public Health, Economics and Health Directorate, Canton of Basel-Landschaft, Switzerland; Department of Biology, Georgetown University, Washington, District of Columbia, USA

**Keywords:** pooled testing, school transmission, COVID-19, retrospective analysis, data-driven contact networks

## Abstract

Pooled testing programs were introduced during the COVID-19 pandemic to expand surveillance capacity while preserving testing resources, but evidence on their epidemiological impact in schools under real-world conditions remains limited.

We analyzed data from the pooled testing program implemented in public primary schools of the canton of Basel-Landschaft, Switzerland, during the Fall-Winter 2021 Delta wave. We used an agent-based transmission model informed by pooled and individual testing results, school characteristics, contact networks, and community incidence. The model was fitted to pooled positivity ratios in four clusters of administrative areas with similar epidemic trajectories. We compared pooled testing with alternative protocols in terms of school transmission, testing volume, and student-days lost.

During the study period, pooled testing was offered to 21’187 students across 62 public primary schools, with high and stable participation across clusters (mean 71–79%). The fitted model reproduced observed pool positivity trends well. Compared with pooled testing, reactive class closure, reactive screening, and symptomatic testing were associated with higher in-school transmission, with excess ranging from 50% to 87%, 63% to 104%, and 72% to 133% across clusters. Weekly individual screening achieved similar reductions in transmission but required 15–25 times more tests. Relaxing class closure after depooling substantially reduced student-days lost without increasing transmission.

Under real-world conditions, pooled testing provided an effective and resource-efficient strategy to reduce SARS-CoV-2 transmission in primary schools. Combining early detection of asymptomatic infections with low testing demands, pooled testing offers a scalable approach to school surveillance and control for pandemic response in educational settings.

## INTRODUCTION

Pooled testing strategies were implemented during the COVID-19 pandemic to screen large populations while preserving limited testing resources (1). These involved a two-step process with a single PCR test conducted on the pooled sample from different individuals, followed by individual tests of positive pool members. This approach was implemented in several settings, including in hospitals in Brazil (2) and care homes in Spain (3), workplaces, schools and care centers in Switzerland (4), the armed forces in the USA (5) and the broader community in Israel (6), Rwanda (7), South Africa (7) and China (8). In educational settings, pooled testing was used in a few instances, including in schools in Germany (9–14), Austria (15), India (16), and in several US schools and universities (17–25), over periods ranging from several weeks (10) to several months (18,20).

In the canton of Basel Landschaft (BL) in Switzerland, a weekly pooled PCR testing program (26) using saliva samples was implemented in all schools of the canton starting in March 2021, replacing the class closure protocol initially implemented after schools reopened in May 2020 to enable face-to-face teaching, while containing transmission risk. In BL, this approach remained in place throughout 2021 and differed from those applied in other cantons after the summer break in 2021. Other cantonal strategies included application of typical contact tracing (Neuchâtel), mask mandates and symptomatic testing with the isolation of positive cases (Vaud) and reactive testing at class level following the confirmation of positive cases in a class, with those testing positive required to isolate (Valais) (27). Similar heterogeneity in school strategies was also observed across Europe (28), highlighting the complex challenges posed by the COVID-19 pandemic. The diversity in approaches often arose from the rapidly evolving nature of the crisis, with each country tailoring its response based on health priorities, available resources, and local circumstances.

While the feasibility of pooled testing programs and their acceptability among students, families and teachers have been widely reported (9–11,20,25), few studies have quantified their epidemiological impact in schools. Evidence from Massachusetts indicated only a modest reduction in school-based infections (22), whereas studies conducted on university campuses reported consistently lower incidence compared to the surrounding community (19,21). Nevertheless, robust evidence on the effectiveness of pooled testing in reducing viral circulation in schools under real-life conditions, particularly in comparison with alternative testing strategies, remains scarce. Moreover, evidence is lacking on the broader consequences of pooled testing, such as the number of student-days lost and the testing resources required. Evaluating these outcomes in the Swiss school context—where pandemic management and educational structures differ from those of previously studied settings— is therefore essential to assess the overall value and applicability of pooled testing approaches.

We used an agent-based transmission model informed by multiple data sources (pooled testing and individual testing results, age-stratified incidence from community surveillance, school records) recorded during the implementation of the pooled testing program to evaluate the effectiveness of the pooled testing program in reducing within school transmission compared to common school-based measures applied simultaneously elsewhere in Switzerland and European countries. We also compared the student-days loss resulting from each measure, reflecting associated disruption to students, as well as the number of tests required, reflecting resource needs.

Our findings go beyond the specific context of SARS-CoV-2, offering valuable guidance for developing proactive strategies in educational settings during future pandemics involving respiratory viruses. This is particularly important in indoor environments where students interact closely (29) and pathogens spread easily (30). This study provides evidence-based insights for a scalable and resource-efficient approach combining early detection, containment and in-person learning.

## METHODS

### Pooling strategy in Basel-Landschaft schools

The pooled testing strategy was initiated in schools with a phased approach, replacing class closures triggered by the identification of positive individuals through symptomatic testing. Starting in week 8 of 2021 (February 22, 2021), schools could implement this strategy on a voluntary basis. By week 12 of 2021 (March 22, 2021), all schools were mandated to offer pooled testing to their students.

Under this strategy, weekly testing consisted of two-stages: in the first stage, saliva samples of individuals (students and teachers) from each class were pooled, and this pool subjected to a PCR test. Results were provided by the end of the same day. In the second stage, classes with a positive pooled test were closed within 24 hours, while pool members underwent individual PCR testing (12-18h turnaround time, depooling step). Positive individuals entered a 10-day isolation, and classes were reopened by decision of a medical board on a case-by-case basis. The protocol applied only to asymptomatic individuals who had not tested positive in the past three months (31). Students showing symptoms underwent individual testing outside schools and isolated for 10 days if positive. Student participation was conditional upon parental consent.

The pooled testing program was paused during school holidays (see Table S2 in the Supplementary Information) and spanned multiple periods, each characterized by a different circulating variant of SARS-CoV-2. Period 1 (weeks 15–26, April 12–July 28, 2021) was dominated by the Alpha variant, which accounted for 87% of sequenced cases in Switzerland-Liechtenstein (32). Period 2 (weeks 33– 39, August 16–October 3, 2021) coincided with the dominance of the Delta variant, responsible for over 99% of sequenced cases (32). Period 3 (weeks 42–50, October 18–December 19, 2021) included a second Delta wave (96% of sequenced cases in weeks 42–50 (32)), and was later followed by the emergence of the Omicron variant (4% of cases (32)).

Although the pooled testing program spanned three epidemic periods (Periods 1–3), our quantitative analysis focused on Period 3 (weeks 42–50, October–December 2021). This period was characterized by the highest infection levels in schools, allowing robust model fitting and inference of transmission dynamics. Earlier periods were used only to inform the temporal clustering of MedStats (see below) to group together areas with consistently similar epidemiological behavior throughout the pandemic. Descriptions of Periods 1 and 2 are provided in the Supplementary Information (SI).

### Databases used in the study and data treatment

The pooling database tracked the number of weekly pooled tests conducted at school, their results, and the corresponding number of participants per MedStat region. MedStat regions are managed by the Federal Statistical Office (FSO) and are designed to analyze healthcare services, specifically to map patient residence while ensuring confidentiality (33). For a MedStat or cluster of MedStats (see section *Inclusion criteria and clustering*), we computed the weekly pool positivity ratio as the fraction of positive pools among tested pools. We used the pool positivity ratio to fit the transmission model (see section *Inference framework)*.

The individual testing database contained the number of tests conducted at testing centers by reasons, including the number and results of depooling tests, and the number of classes closed per week and MedStat. We harmonized the pooling and individual testing databases to ensure a one-to-one correspondence between positive pools and their associated depooling results, thereby removing inconsistencies such as positive pools without a corresponding depooling entry or vice versa (see SI for details on the cleaning procedure). We used the observed pool positivity ratio together with the corresponding fraction of positive students at depooling to initialize the model with a certain number of positive students (see section *Simulations*). In addition, we combined the number of closed classes with the number of positive pools (pooling database) to inform the model with the probability of class closure associated with a positive pool (see SI).

Community surveillance data on weekly case incidence for primary school-aged children (5-12 years old) and adults (18-59 years old) at the MedStat level were used to cluster MedStats (see *Inclusion criteria and clustering)* and to estimate infection events outside school settings in each cluster.

Incidence was determined from individual tests, independently of the testing reason. Estimated infection events outside schools were used to introduce positive cases throughout the simulations (see section *Simulations*).

Data on the weekly proportion of fully vaccinated individuals (defined as prior infection plus one dose, or two doses) and boosted individuals (fully vaccinated with an additional dose) obtained from the Cantonal Office of Public Health were used to inform vaccination rates in the school population.

The school database recorded the number of schools, number of classes and number of students per grade level, as well as the median class size per grade level. We used the number of students per grade, and number of classes per grade to retrieve school population characteristics by MedStat cluster and generate the corresponding contact networks to inform transmission in our model. Moreover, combining the individual testing database and the school database, we obtained the number of students present per week per MedStat as the number of students not in isolation. We combined this with the number of pool participants from the pooling database to compute the weekly participation rate to the pooling program, which was also integrated in our model.

Finally, we used data on communal socio-economic characteristics and rural-urban classification (34,35) to characterise the different clusters of MedStats identified (see section *Inclusion criteria and clustering*). We characterised them in terms of the fraction of residents in rural, intermediate and urban communes and the public social welfare expenses per inhabitant.

All accessed information was anonymized and aggregated by MedStats within the BL canton.

### Inclusion criteria and clustering

To reduce bias due to small student sample size, we considered MedStats with at least 500 registered students (Criterion #0). Then, in order to identify MedStats with similar epidemic profiles, we used an agglomerative hierarchical clustering algorithm (36) to group MedStats based on pairwise correlations in weekly case incidence in children aged 5-12 years old, between weeks 15 and 50 of 2021 (12/04/2021 – 19/12/2021). MedStats were merged according to progressively weaker correlations, such that MedStats within the same cluster followed mutually similar temporal epidemic patterns.

To ensure that the modelling analysis was based on pooled testing data with adequate participation, we included, for each cluster and week, only those pooled testing results where student participation reached at least 60% (Criterion #1). To ensure frequent pooling implementation, we furthermore only included MedStats where at least four pooled testing results were available (Criterion #2) in the period under study (Period 3).

For sensitivity, we assessed the robustness of the clustering analysis to the definition of Criterion #0 by considering MedStats with at least 300, or at least 750 registered students. In addition, we explored different participation levels for Criterion #1 by setting the threshold to 50%, or 75% to assess their impact on pooled testing effectiveness.

### Data-driven temporal school contact networks

For each identified MedStat cluster, we constructed a data-driven temporal school network. In each network, nodes correspond to children or teachers and links between them represent the occurrence of a face-to-face contact of at least 20 seconds. We used high-resolution empirical data on face-to-face proximity interactions gathered in a French primary school in 2009 (29) to infer patterns of contact behavior and adapt them to the Swiss schools under study to generate links between students. For each cluster, we adjusted the size and class structure in the contact data (29) to match the average school size, the average number of students per class, and number of classes per grade observed in each cluster, as described in the SI. We also adjusted the temporal networks in order to account for the Swiss school system’s schedule and structure (37). This involved timetable changes, and the addition of grades for kindergarten and grade 8 students, absent in the empirical contact data set (see the SI). As the empirical contact data only covered two days and looping over them is known to bias epidemic outputs (38), we extended the contact networks to match the complete study period with an algorithm previously formulated to synthetically extend school contact networks over successive days (38).

### Transmission model

We extended a stochastic agent-based model originally developed to study SARS-CoV-2 transmission in schools during the Alpha, Delta, and Omicron waves in France (39–42). Interactions in the school population were encoded in the data-driven contact networks obtained for each cluster (see section *Data-driven temporal school contact networks*). Transmission occurred with transmissibility rate *β*per contact per unit time when a susceptible and infectious individual were in contact. The transmissibility rate *β* was fitted to reproduce the pool positivity ratio estimated in each cluster (see section *Inference Framework*). Infection progression included prodromic transmission followed by a clinical or subclinical disease stage, informed by empirical distributions (43–45). We included a transient phase before fully recovering, where individuals were no longer infectious but remained detectable by PCR test. The model was parametrized with age-specific estimates of relative susceptibility, infectiousness, probability of developing a clinical infection, and probability of case detection based on symptoms (46–51); and it was adapted to account for the Delta variant-specific epidemiological and immunological characteristics (52). Individuals were stratified by vaccination status, distinguishing fully vaccinated individuals from those boosted with an additional dose. Age-specific vaccination coverage was informed by the Cantonal Office of Public Health, while sero-prevalence estimates were used to determine initial immunity levels (53,54). We implemented age-specific vaccine effectiveness against infection, transmission, and development of symptoms following infection, depending on vaccination status (55). Thorough details on the model structure and parameters can be found in the SI.

### Simulations

We simulated the pooled testing protocol in the school population constructed in each cluster. Simulations started at the beginning of Period 3, when students returned to school after a break during which transmission chains at school were interrupted.

We initialized simulations with positive students derived from the prevalence observed at depooling in positive pools. Weekly introductions of exposed individuals were estimated stochastically using case incidence data. Because case incidence reflects infections acquired both inside and outside schools, the number of introductions was adjusted during simulations to account for infections occurring within the school setting, as in previous work (39). Further details are provided in the SI.

We simulated the program on a fixed weekly schedule, with pooled tests conducted on Mondays. On Tuesdays, positive classes were closed, while individual PCR tests were simulated for depooling. As the decision to reopen each class was taken case-by-case without consistent guidelines, we estimated the weekly rolling probability of class closure following a positive pool result per cluster of MedStats from the data and simulated 5-day closures accordingly (see SI). The participation rate to the pooled testing each week was informed by the weekly participation measured in each cluster. When simulating a pooled test, we estimated for each pool the probability of the test result being positive from the number of infected participants participating to the pool, their disease stage (4,56), and the pooled test sensitivity (see SI). Individuals with a clinical infection may also be identified with a PCR test (symptom-based testing). Positive individuals identified at depooling or from symptoms all entered a mandated 10-day isolation period.

We assumed 8h turnaround times for all PCR tests. We assumed pooled tests sensitivity and specificity of 75% and 100% respectively (57,58). Individual test sensitivity of depooling tests, and under the alternative strategies considered (see section *Effectiveness of pooled testing*) was instead simulated time-varying, peaking at 96% (59). For sensitivity analysis, we also explored values of pooled tests sensitivity fixed to 65% and 85%.

### Effectiveness of pooled testing

We first evaluated the benefit of the pooled testing protocol in comparison to the previously implemented reactive class closure protocol. This protocol consisted of 10-day class closures when a positive case was identified through symptom-based testing. We also compared the pooled testing strategy with weekly individual screening, with adherence in line with participation rates observed during the pooled testing program, to understand the added value of pooling the tests. Finally, we compared the pooled testing protocol to two other school-based protocols applied in other Swiss cantons (27). These involved the reactive individual screening protocol (Valais), and the symptoms-based testing protocol (Vaud). The reactive individual screening protocol aimed to avoid widespread class closures by requiring classmates of detected cases to test themselves. We simulated reactive screenings on days 0, 2, and 4 after the initial detection, with adherence in line with participation to the pooled testing program and 10-day isolation of non-compliant students. The symptom-based testing protocol involved only the sole testing of symptomatic individuals, with the 10-day isolation of positive individuals. To ensure comparability, all alternative protocols were simulated under the same epidemic conditions estimated for the pooled testing. Specifically, we seeded the alternative protocols with the same weekly introductions estimated under the pooled testing and applied the same estimated transmissibility rate. Individual tests in all protocols were simulated as salivary PCR tests, as in the pooled testing protocol.

We evaluated the performance of the pooled testing protocol by assessing differences in school transmission, testing volume, and student-days lost with alternative testing strategies. Excess school transmissions for each alternative protocol were defined as the difference between the number of school-based transmissions observed under the alternative protocol and those observed under the pooled testing, expressed relative to the pooled testing baseline. The increase in testing volume for regular testing protocols was computed as the n-fold increase of the cumulative number of individual tests required compared with pooled testing. The fraction of student-days lost was computed for each protocol as the cumulative number of student-days spent in isolation or quarantine over the total number of student-days.

### Inference framework

We estimated the within-school transmissibility per contact per unit time (*β*/min) together with a scaling parameter *f* adjusting the number of community introductions derived from surveillance incidence. Parameters were estimated using a maximum likelihood estimation approach by fitting the model to the pool positivity ratios in each cluster. Pairs of *β* and *f* were estimated by grid exploration at the maximum likelihood, assuming a binomial distribution for the number of positive pools among those tested. Estimates were obtained for each cluster from 1,000 simulated stochastic outbreaks for each parameter pair, considering time-varying participation and class closure probability observed in each cluster. Further details on the inference procedure are provided in the SI.

## RESULTS

### Pooling program overview in Basel-Landschaft canton

In Period 3, pooled testing was offered to 21,187 students, across 62 public primary schools (grades 1-8, ages 5-12 years old) in the 21 (out of 23) MedStats meeting Criterion #0. The application of Criterion #1 to weekly pooled testing results excluded 1 MedStat in weeks 45 and 50. All data entries meeting Criterion#0 and #1 also met Criterion #2. A total of 10,817 pools were considered after accounting for the inclusion criteria (representing 94% of all pools). Among these, 455 (4.2%) were positive throughout the study period. Student positivity rate at depooling was 12.0%. As 1,385 individual tests were performed additionally on 5-12 years old children in parallel to the school pooled testing scheme, depooling tests represented 82.2% of the individual testing effort in the child population.

### Clustering analysis

We identified four distinct clusters of MedStats, based on individual case incidence in children of 5 to 12 years old (**Figure 1A**). Each cluster varied in the number of MedStats included (**Figure 1B**). Cluster 2 included the highest number of schools and students (38 schools and 12,863 students) followed by Cluster 4 (15 schools and 3,704 students), Cluster 3 (6 schools and 2,165 students) and Cluster 1 (3 schools and 2,455 students).

**Figure 1.**
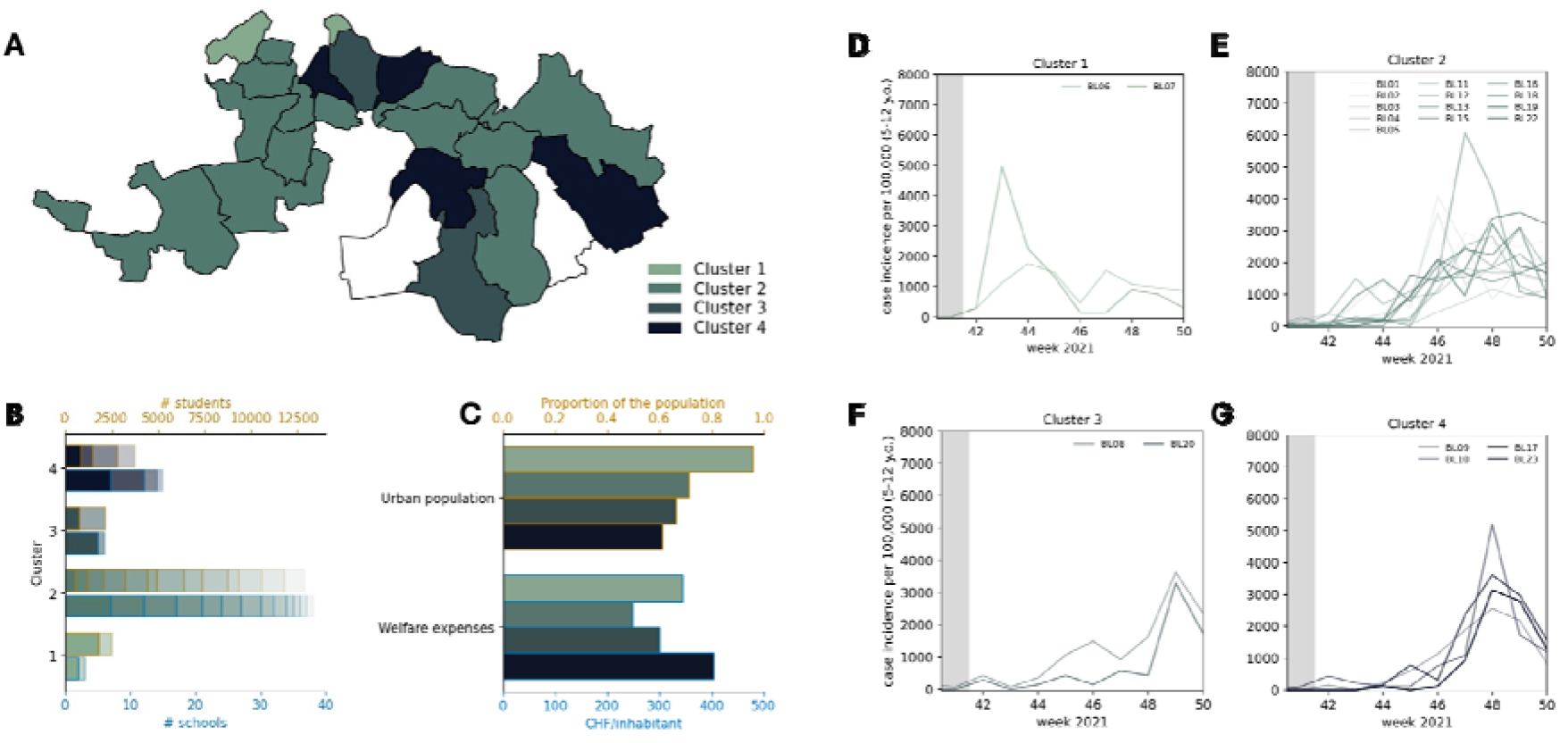
Characterization of the Clusters of MedStats identified from children incidence curves. **A** Map of the MedStats in the canton of Basel-Landschaft, coloured by cluster membership (white MedStats are those not meeting inclusion criterion #0). **B** Number of schools and number of students in each cluster, with each shade layer corresponding to a different MedStat. **C** Proportion of the population living in urban communes and public expenses in social welfare per inhabitant for each cluster. **D** Incidence curves in children 5-12 years old in Cluster 1 as identified by the hierarchical algorithm. **E** As in D for Cluster 2. **F** As in D for Cluster 3. **G** As in D for Cluster 4.

Our analysis of the socio-economic characteristics of the clusters (**Figure 1C**) identified Cluster 1 as predominantly urban, with 96% of its population residing in urban communes. Clusters 2, 3 and 4 were more heterogeneous, with 71%, 66%, and 61% of residents living in urban areas, respectively. Public social welfare support was highest in Cluster 4, reaching 403 CHF per inhabitant, compared with 344, 301 and 248 CHF per inhabitant in clusters 1, 3, and 2 respectively.

The clustering grouped MedStats with similar temporal patterns of SARS-CoV-2 incidence in children aged 5–12 years (**Figure 1D–G**). Cluster 1 exhibited an earlier epidemic peak around week 43–44. Cluster 2 and 4 showed a later wave peaking around week 47-48, with incidence increasing steadily in Cluster 2 and showing a sharper rise and decline in Cluster 4. Clusters 3 experienced a later peak, around week 49.

### Participation and pooled testing effort by clusters during the fall-winter Delta wave

Participation in the pooled testing program remained consistently high across all clusters, with mean participation rates equal to 78% in Cluster 1, 79% in Cluster 2, 77% in Cluster 3, and 71% in Cluster 4 (**Figure 2A**). The weekly number of participating students and pools varied by cluster due to differences in sizes (**Figure 2B** and **2C**), averaging 9,649 students and 739 pools per week in Cluster 2, 1,846 students and 139 pools per week in Cluster 1, 1,596 students and 119 pools per week in Cluster 3, and 2,335 students and 205 pools per week in Cluster 4. Both the number of participating students and of pools tested remained stable over the period, except for decreases in weeks 45 and 50, when specific MedStat records were excluded following Criterion #1. Overall, 99.5% of pooled tests included in the analysis had student participation above 60%. The distribution of participation rates in all included MedStats is provided in the SI.

**Figure 2.**
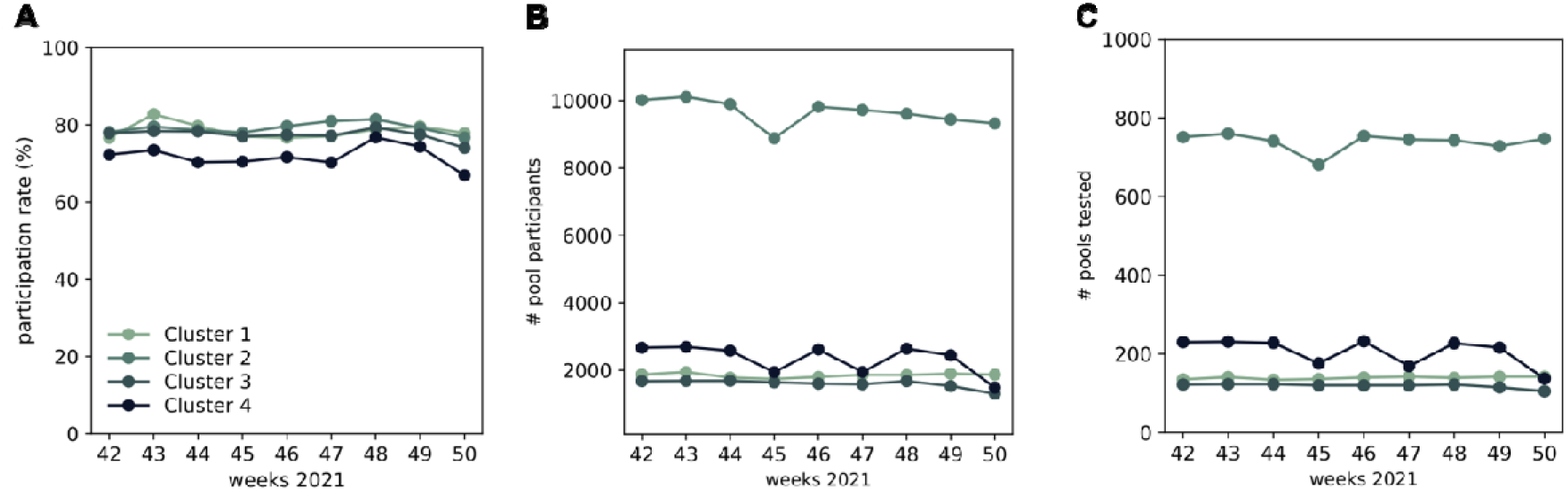
Pooling strategy by clusters during the fall peak in 2021. **A**. Weekly participation in the pooled testing program **B**. Weekly number of participants providing a sample for pooled testing **C**. Weekly number of pools tested.

### Pool positivity ratio

The weekly pool positivity ratio predicted by the fitted models closely matched the observed data in the four clusters (**Figure 3A-D**) and reproduced the main temporal patterns observed in incidence (**Figure1D-G**). In Clusters 1 and 4, the model captured both the timing and magnitude of the epidemic waves (**Figure 3A, D**). In Cluster 2, the timing of the peak was reproduced, although its magnitude was slightly overestimated (**Figure 3B**). In Cluster 3, the model captured well the timing and magnitude of the peak around week 49, with a slight mismatch in the decline. These results were robust to alternative assumptions on pooled test sensitivity (see SI).

**Figure 3.**
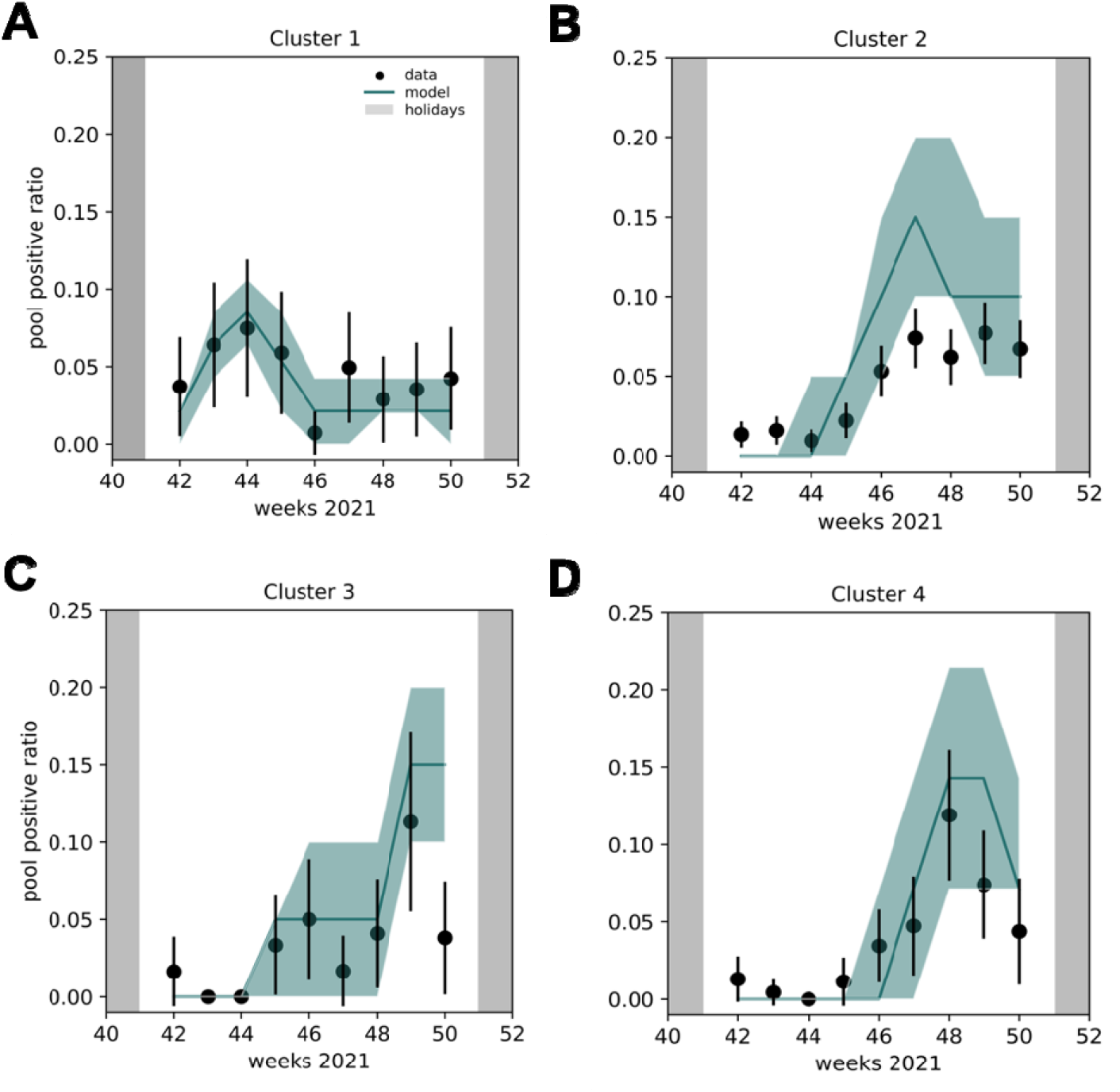
Pool positivity ratio observed in the identified clusters of MedStats during the fall Delta wave in 2021, together with model estimates. **A** The weekly pool positivity ratio estimated by the model in Cluster 1 (green line) is compared to the observed pool positivity ratios (black dots). The green area represents the interquartile range (IQR), and the error bars encode 95% confidence intervals. **B** As in A for Cluster 2. **C** As in A for Cluster 3. **D** As in A for Cluster 4. Model estimates in all panels are obtained from 1,000 numerical simulations. In all panels, pool positivity ratio is defined as the fraction of positive pools among tested pools.

The fitted model also provided estimates of cluster-level epidemic outcomes. Median attack rates ranged from 15.1% to 38.2% across clusters, and school-acquired infections accounted for between 50.0% and 71.8% of total infections (see SI).

### Effectiveness of the pooling strategy

Across all clusters, symptomatic testing, reactive class-closure, and reactive screening strategies were predicted to result in substantially higher in-school transmission compared with the pooled testing protocol (**Figure 4A–D**). Reactive class closure—the strategy replaced by pooled testing in February 2021—was associated with excess in-school transmission ranging from 50% (IQR 0–107%) in Cluster 4 to 87% (IQR 33-148%) in Cluster 2 compared to pooled testing. Reactive screening produced similar but slightly higher increases, with excess transmission between 63% (IQR 16-126%) in Cluster 4 and 104% (IQR 44-174%) in Cluster 2. Symptomatic screening, as implemented in the canton of Vaud, produced the largest increases, ranging from 72% (IQR 28-133%) in Cluster 1 and 133% (IQR 70-204%) in Cluster 2. The weekly individual screening protocol achieved the lowest transmission levels among the alternative strategies, with values close to those of pooled testing. However, it required substantially more resources, with an estimated 15-to 25-fold increase in the number of tests across clusters (**Figure 4E**). Relaxing the class closure after depooling during pooled testing instead did not lead to significant excess transmission compared to the baseline protocol involving case-by-case class closure after positive depooling.

**Figure 4.**
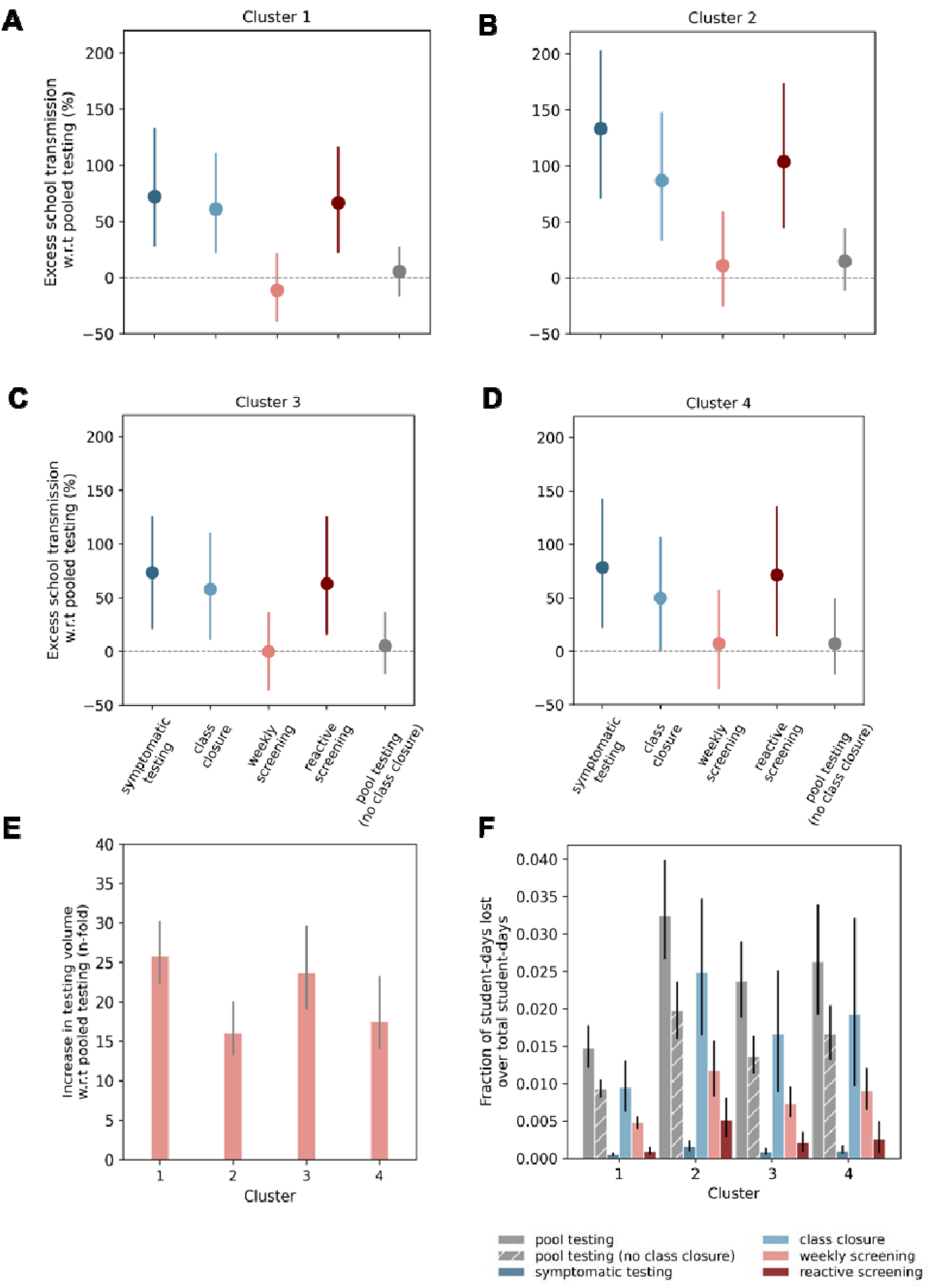
Comparison between the pooled strategy and alternative school protocols. **A**. Percentage of school transmissions in excess with the symptomatic screening, class closure, weekly individual screenings and reactive screening with respect to the pooled testing protocol in Cluster 1. **B**. Same as A for Cluster 2. **C**. Same as A for Cluster 3. **D**. Same as A for Cluster 4. **E**. Increase in testing volume of the weekly individual screening protocol compared to the pooling strategy **F**. Fraction of student-days lost over the total number of student-days under the class pooled testing, symptomatic testing, class closure, weekly and reactive screening protocols with adherence as observed during the pooled testing.

The pooled testing protocol was estimated to cause the largest fraction of student-days lost in all four clusters (**Figure 4F**), when implemented with systematic class closure between positive pooling and depooling, and with case-by-case class closure after depooling. Under this configuration, the fraction of student-days lost ranged between 1.5% (IQR 1.2-1.8%) in Cluster 1 and 3.2% (IQR 2.6% - 4.0%) in Cluster 2. Relaxing the second type of class closure after depooling substantially reduced these losses, lowering the fraction of student-days lost to between 0.9% (IQR 0.8–1.1%) in Cluster 1 and 2.0% (IQR 1.6–2.4%) in Cluster 2, without generating significant excess transmission relative to the baseline pooled testing protocol. Under this modified protocol, student-days lost were also lower than those associated with the reactive class-closure strategy in three of the four clusters.

For comparison, the reactive class closure protocol previously in place in the canton resulted in between 1.0% (IQR 0.6-1.3%) and 2.5% (IQR 1.6% −3.5%) of student-days lost in Cluster 1 and in Cluster 2, respectively. Symptomatic testing alone produced the lowest disruption, with fractions ranging from 0.05% (IQR 0.03-0.08%) in Cluster 1 and 0.2% (IQR 0.1-0.2%) in Cluster 2. Weekly individual screening resulted in intermediate levels, between 0.5% (IQR 0.4-0.6%) and 1.2% (IQR 0.8% - 1.6%) of student-days lost in Clusters 1 and 2, respectively.

These results were robust to changes in pool testing sensitivity, and changes in contact rates of the kindergarten classes with the rest of the school, in all clusters (see SI).

## DISCUSSION

This study evaluated the performance of Switzerland’s pooled testing program for SARS-CoV-2 in schools in the canton of Basel-Landschaft during the Fall-Winter 2021 Delta wave. Using an agent-based model calibrated to pooled testing data, we compared the observed strategy with alternative school testing protocols under the same epidemic conditions. Pooled testing consistently reduced in-school transmission compared with symptomatic testing, reactive screening, and reactive class-closure strategies across all clusters. While weekly individual testing achieved similar epidemiological performance, it required substantially more testing resources. Although pooled testing with temporary class closure generated higher student-days lost than some alternatives, relaxing class closure after depooling substantially reduced this burden without increasing transmission. Participation in the program remained high and stable across clusters, supporting the feasibility of large-scale implementation. Together, these findings indicate that pooled testing provides an effective and resource-efficient approach for monitoring and mitigating SARS-CoV-2 transmission in school settings under real-world conditions.

Our findings provide quantitative evidence that pooled testing can reduce in-school SARS-CoV-2 transmission compared with reactive strategies. Regular screening of students through pooled sampling enables earlier detection of asymptomatic infections, thereby interrupting transmission chains before they expand within schools. In contrast, reactive strategies based on symptom detection or contact tracing intervene only after cases are identified, allowing undetected spread to occur before control measures are implemented (39,40,42,60,61). Consistent with this mechanism, pooled testing was predicted to avert school transmission more effectively than reactive class closure (excess transmission 50-87%), reactive screening (63-126%), and symptomatic testing (72-133%). Quantitative estimates of the epidemiological impact of pooled testing remain scarce. Observational studies from US schools and universities have reported lower infection incidence in settings implementing pooled testing compared with surrounding communities. A pooling program found a consistently lower (50%-78%) positivity rate in schools undergoing pooling tests than in the general community (18). Similarly, the Duke university pooled testing program was associated with a weekly average per-capita infection incidence of 0.08% on campus, compared to 0.1% in the community (19). At Georgia Institute of Technology, pooled testing results showed a 3-5 fold reduction in incident cases compared to the full county (21). However, comparisons across settings are complicated by differences in community infection pressure and non-pharmaceutical interventions mandated at the same time in the community (62). Our analysis contributes quantitative evidence by explicitly comparing school transmission estimated under pooled testing with that expected under alternative protocols under the same epidemic conditions. These results strengthen evidence that pooled testing can effectively reduce SARS-CoV-2 transmission while maintaining continuity in educational settings.

Pooled testing also offers a major advantage in testing efficiency compared with weekly individual screening. In our analysis, both strategies achieved similar reductions in in-school transmission but required markedly different testing volumes: weekly individual screening would have required up to 25 times more PCR tests than pooled testing. This difference has important implications for feasibility, and the large testing volume required by individual screening is often cited as a barrier to large-scale implementation. By combining multiple samples into a single test and depooling only positive pools, pooled testing substantially reduces the number of diagnostic tests needed while maintaining epidemiological effectiveness. The efficiency of pooling depends on infection prevalence and pool composition. Previous modeling studies have shown that when prevalence is at 5% and the reproductive number reaches 3, both strategies could require the same volume of tests when pools are not formed by shared exposure environments (i.e. classrooms) (63). Instead, by pooling together individuals likely to be infected concurrently, e.g. classmates, office mates or individuals close socially, the amount of tests required by pooled strategies diminishes (18,63,64), as fewer pools need depooling. Our analysis shows that under the epidemic conditions observed during the Fall-Winter Delta wave of 2021 in Switzerland, the classroom-based pooled testing strategy provided a substantial reduction in testing resources. Several studies of pooled testing programs in educational environment have similarly highlighted their economic advantages compared to individual tests (16,19,24,63). Nevertheless, the cost of these programs remains high, particularly in settings with limited resources (25). These costs should be considered alongside the potential economic burden associated with uncontrolled outbreaks when less effective protocols are implemented (65).

School testing strategies must also be evaluated in terms of their impact on educational continuity. In our study, the pooled testing program implemented with case-by-case class closure after depooling resulted in slightly more student-days lost compared to the previously applied reactive class closure protocol across clusters. This effect was most pronounced in Cluster 2, which also showed a higher estimated contribution of in-school transmission and a larger fraction of students infected (see SI). Relaxing class closure after depooling substantially reduced this burden, resulting in fewer student-days lost, while maintaining similar transmission control. Weekly individual screening would have led to fewer student-days lost than pooled testing with class closure, largely because students do not need to be temporarily excluded while waiting for depooling results. Symptomatic testing alone would have minimized disruption to in-person learning, but at the cost of substantially higher in-school transmission. More broadly, regular screening strategies may lead to increased isolation due to the identification of asymptomatic infections (61). However, by reducing transmission within schools, pooled testing can also limit the number of cases requiring isolation over time. This mechanism was already observed for weekly individual screening (42). Previous pilot programs (25) have similarly shown that pooled testing can be implemented without increasing school absences and may even reduce them by reassuring students and families about the safety of in-person learning (20).

Participation in the pooling program remained high throughout the study period, approximately 8 months after the start of the protocol implementation, with no evidence of decline over time. Average participation in the four clusters of MedStats ranged between 71% and 79%, and 99.5% of the pooled tests recorded participation above 60%. Participation in Cluster 4 was slightly lower than in the other clusters, which may reflect differences in socio-economic characteristics. In particular, the lower proportion of urban residents in this cluster is consistent with previous evidence reporting lower adoption of COVID-19 protective behaviors in more rural populations (66). More broadly, participation in pooled testing programs has been reported to vary across contexts, with rates typically ranging between approximately 50% and over 90% in school (9–12) and university (19,21) settings. In Germany, participation rates varied between 54% in a feasibility study in two schools (11), 70% in a program covering 14 schools (12), and 92% in a pilot study in 10 primary schools (9). In the US, participation rates varied between 61% and 81% in the same school district (25). Such differences may reflect socio-economic, cultural, and political factors that influence the adoption of preventive behaviors, including urbanicity, education, and trust in institutions (66–69). Because participation in school testing programs often depends on parental consent, these factors may also indirectly shape participation rates among children.

Our study has several limitations. First, the contact networks used in our model were constructed synthetically from empirical contact data gathered in a French primary school in 2009 and may therefore not fully represent contact patterns in Swiss primary schools. Collecting empirical contact data in schools is costly and logistically and ethically challenging as parental consent is required. While contact data have recently been collected in Swiss high schools (70), these datasets do not capture interactions among primary school children. To mitigate this limitation, we constructed synthetic contacts to reflect key features of the Swiss school system, including timetable structure, grade composition, and reduced mixing with kindergarten classes. Importantly, our results were robust to alternative assumptions used to reconstruct these networks. Second, the model focused on transmission occurring within schools and did not explicitly represent transmission in households or the broader community. However, we accounted for all transmission events outside school by estimating the weekly number of introductions (39). Third, our findings were derived for primary school settings and may not be directly generalizable to high-school or university settings where student density, contact behaviors, timetables, as well as epidemiological and immunological characteristics differ. Observational studies in secondary schools and university campuses have reported pooled testing lowering infection incidence compared with surrounding communities (18,19,21), but they did not quantify infections averted or directly compare alternative testing strategies. Further work is needed to assess the effectiveness of pooled testing protocols in these contexts. Finally, our analysis was conducted during the Delta wave of SARS-CoV-2 and under the control measures implemented in Switzerland at that time. Changes in viral transmissibility, population immunity, or testing protocols—such as those introduced during the subsequent Omicron wave—may affect both the effectiveness of testing strategies and the resources required to implement them (62). Previous work has shown that weekly individual screening remains effective even at high incidence while requiring testing resources comparable to reactive strategies (40). Pooled testing is expected to remain similarly effective compared to other strategies, although its resource advantage may diminish as incidence increases because a larger fraction of pools requires depooling.

These findings support pooled testing as a scalable and resource-efficient surveillance strategy for schools. By enabling early identification of infections, including asymptomatic cases, pooled testing can help limit in-school transmission while avoiding the large testing demands associated with individual screening. At the same time, the results highlight the importance of carefully designing isolation and closure policies accompanying testing programs to minimize educational disruption. Together, these elements suggest that pooled testing can represent a sustainable approach for maintaining safe in-person education during periods of elevated respiratory virus circulation.

Overall, pooled testing offered the most balanced strategy among those evaluated, combining effective control of in-school transmission with efficient use of testing resources and manageable educational disruption. These results provide empirical support for the role of pooled testing as a practical surveillance tool in schools and underline its potential value for managing future infectious disease outbreaks in educational settings.

## Contributors

V.C. conceived and designed the study. J.A.B. was involved in the pooled testing program. K.N.K. preprocessed data. E.C. and L.C. analyzed the data. E.C. developed the code and ran the simulations. All authors interpreted the results. E.C., L.C. and V.C. wrote the initial manuscript draft. All authors edited and approved the final version of the Article.

## Ethics approval

A request for clarification of responsibility under the Human Research Act was submitted to the EKNZ (Ethikkommission Nordwest und Zentralschweiz) and was deemed not to require formal ethical approval with no issues of concern identified (Req-2022-00005).

## Data sharing Statement

The data that support the findings of this study will be made available upon reasonable request for scientific research purposes. For further information or to request access, please contact Katrin N. Koch or Julia A. Bielicki.

## AI disclosure

ChatGPT (OpenAI) and LeChat (Mistral AI) were used for minor text editing, and the suggested modifications were carefully reviewed by EC and LC. All authors reviewed the manuscript after AI use and take full responsibility for the material submitted.

## Acknowledgments

We thank Anthony Hauser for helpful discussions.

## REFERENCES

1. Grobe N, Cherif A, Wang X, Dong Z, Kotanko P. Sample pooling: burden or solution? Clinical Microbiology and Infection. 2021 Sep;27(9):1212–20. doi:10.1016/j.cmi.2021.04.007

2. Christoff AP, Cruz GNF, Sereia AFR, Boberg DR, Bastiani DC de, Yamanaka LE, et al. Swab pooling: A new method for large-scale RT-qPCR screening of SARS-CoV-2 avoiding sample dilution. PLOS ONE. 2021 Feb 4;16(2):e0246544. doi:10.1371/journal.pone.0246544

3. Cabrera Alvargonzalez JJ, Rey Cao S, Pérez Castro S, Martinez Lamas L, Cores Calvo O, Torres Piñon J, et al. Pooling for SARS-CoV-2 control in care institutions. BMC Infectious Diseases. 2020 Oct 12;20(1):745. doi:10.1186/s12879-020-05446-0

4. Riou J, Studer E, Fesser A, Schuster TM, Low N, Egger M, et al. Surveillance of SARS-CoV-2 prevalence from repeated pooled testing: application to Swiss routine data. Epidemiol Infect. 2024 Aug 22;152:e100. doi:10.1017/S0950268824000876 PubMed PMID: 39168632.

5. McGhee LL, Yerramilli SV, Nadolny R, Casal C, Kearney BM, Taylor H, et al. Implementing Pool-Based Surveillance Testing for SARS-CoV-2 at the Army Public Health Center Laboratory and across the Army Public Health Laboratory Enterprise. Med J (Ft Sam Houst Tex). 2021 Jan 1;(PB 8-21-01/02/03):83–9. PubMed PMID: 33666917.

6. Barak N, Ben-Ami R, Sido T, Perri A, Shtoyer A, Rivkin M, et al. Lessons from applied large-scale pooling of 133,816 SARS-CoV-2 RT-PCR tests. Science Translational Medicine. 2021 Apr 14;13(589):eabf2823. doi:10.1126/scitranslmed.abf2823

7. Mutesa L, Ndishimye P, Butera Y, Souopgui J, Uwineza A, Rutayisire R, et al. A pooled testing strategy for identifying SARS-CoV-2 at low prevalence. Nature. 2021 Jan;589(7841):276–80. doi:10.1038/s41586-020-2885-5

8. Cao S, Gan Y, Wang C, Bachmann M, Wei S, Gong J, et al. Post-lockdown SARS-CoV-2 nucleic acid screening in nearly ten million residents of Wuhan, China. Nat Commun. 2020 Nov 20;11(1):5917. doi:10.1038/s41467-020-19802-w

9. Kästner A, Lücker P, Sombetzki M, Ehmke M, Koslowski N, Mittmann S, et al. SARS-CoV-2 surveillance by RT-qPCR-based pool testing of saliva swabs (lollipop method) at primary and special schools-A pilot study on feasibility and acceptability. PLoS One. 2022;17(9):e0274545. doi:10.1371/journal.pone.0274545 PubMed PMID: 36099277; PubMed Central PMCID: PMC9469960.

10. Kretschmer AC, Junker L, Dewald F, Linne V, Hennen L, Horemheb-Rubio G, et al. Implementing the Lolli-Method and pooled RT-qPCR testing for SARS-CoV-2 surveillance in schools: a pilot project. Infection. 2023 Apr;51(2):459–64. doi:10.1007/s15010-022-01865-0 PubMed PMID: 35759174; PubMed Central PMCID: PMC9243733.

11. Sweeney-Reed CM, Wolff D, Niggel J, Kabesch M, Apfelbacher C. Pool Testing as a Strategy for Prevention of SARS-CoV-2 Outbreaks in Schools: Protocol for a Feasibility Study. JMIR Res Protoc. 2021 May 28;10(5):e28673. doi:10.2196/28673 PubMed PMID: 33979297; PubMed Central PMCID: PMC8166266.

12. Joachim A, Dewald F, Suárez I, Zemlin M, Lang I, Stutz R, et al. Pooled RT-qPCR testing for SARS-CoV-2 surveillance in schools - a cluster randomised trial. EClinicalMedicine. 2021 Sep;39:101082. doi:10.1016/j.eclinm.2021.101082

13. Gruendl M, Kheiroddin P, Althammer M, Schöberl P, Rohrmanstorfer R, Wallerstorfer D, et al. Analysis of COVID-19 Infection Chains in a School Setting: Data From a School-Based rRT-PCR– Gargle Pool Test System. Disaster Med Public Health Prep. 17:e312. doi:10.1017/dmp.2022.279 PubMed PMID: 36789767; PubMed Central PMCID: PMC9947041.

14. Dewald F, Suárez I, Johnen R, Grossbach J, Moran-Tovar R, Steger G, et al. Effective high-throughput RT-qPCR screening for SARS-CoV-2 infections in children. Nat Commun. 2022 Jun 25;13(1):3640. doi:10.1038/s41467-022-30664-2

15. Willeit P, Krause R, Lamprecht B, Berghold A, Hanson B, Stelzl E, et al. Prevalence of RT-qPCR-detected SARS-CoV-2 infection at schools: First results from the Austrian School-SARS-CoV-2 prospective cohort study. Lancet Reg Health Eur. 2021 Mar 23;5:100086. doi:10.1016/j.lanepe.2021.100086 PubMed PMID: 34396360; PubMed Central PMCID: PMC8350968.

16. Singh AK, Nema RK, Joshi A, Shankar P, Nema S, Raghuwanshi A, et al. Evaluation of pooled sample analysis strategy in expediting case detection in areas with emerging outbreaks of COVID-19: A pilot study. PLOS ONE. 2020 Sep 22;15(9):e0239492. doi:10.1371/journal.pone.0239492

17. Simas AM, Crott JW, Sedore C, Rohrbach A, Monaco AP, Gabriel SB, et al. Pooling for SARS-CoV2 Surveillance: Validation and Strategy for Implementation in K-12 Schools. Front Public Health. 2021 Dec 17;9. doi:10.3389/fpubh.2021.789402

18. Mendoza RP, Bi C, Cheng HT, Gabutan E, Pagaspas GJ, Khan N, et al. Implementation of a pooled surveillance testing program for asymptomatic SARS-CoV-2 infections in K-12 schools and universities. eClinicalMedicine. 2021 Aug 1;38. doi:10.1016/j.eclinm.2021.101028 PubMed PMID: 34308321.

19. Denny TN. Implementation of a Pooled Surveillance Testing Program for Asymptomatic SARS-CoV-2 Infections on a College Campus — Duke University, Durham, North Carolina, August 2– October 11, 2020. MMWR Morb Mortal Wkly Rep. 2020;69. doi:10.15585/mmwr.mm6946e1

20. Berke EM, Newman LM, Jemsby S, Hyde B, Bhalla N, Sheils NE, et al. Pooling in a Pod: A Strategy for COVID-19 Testing to Facilitate a Safe Return to School. Public Health Rep. 2021 Nov 1;136(6):663–70. doi:10.1177/00333549211045816

21. Gibson G, Weitz JS, Shannon MP, Holton B, Bryksin A, Liu B, et al. Surveillance-to-Diagnostic Testing Program for Asymptomatic SARS-CoV-2 Infections on a Large, Urban Campus in Fall 2020. Epidemiology. 2022 Mar;33(2):209. doi:10.1097/EDE.0000000000001448

22. Branch-Elliman W, Ertem MZ, Nelson RE, Danesharasteh A, Berlin D, Fisher L, et al. Impacts of testing and immunity acquired through vaccination and infection on covid-19 cases in Massachusetts elementary and secondary students. Commun Med. 2024 Oct 16;4(1):1–10. doi:10.1038/s43856-024-00619-3

23. Pollock NR, Berlin D, Smole SC, Madoff LC, Brown C, Henderson K, et al. Implementation of SARS-CoV2 Screening in K–12 Schools Using In-School Pooled Molecular Testing and Deconvolution by Rapid Antigen Test. J Clin Microbiol. 59(9):e01123–21. doi:10.1128/JCM.01123-21 PubMed PMID: 34191585; PubMed Central PMCID: PMC8373013.

24. Perea S, Tretina K, O’Donnell KN, Love R, Bethlendy G, Wirtz M, et al. Saliva-Based, COVID-19 RT-PCR Pooled Screening Strategy to Keep Schools Open. Disaster Medicine and Public Health Preparedness. 2023 Jan;17:e70. doi:10.1017/dmp.2021.337

25. Doron S, Ingalls RR, Beauchamp A, Boehm JS, Boucher HW, Chow LH, et al. Weekly SARS-CoV-2 screening of asymptomatic kindergarten to grade 12 students and staff helps inform strategies for safer in-person learning. Cell Reports Medicine. 2021 Nov 16;2(11):100452. doi:10.1016/j.xcrm.2021.100452

26. Projekt «Breites Testen Baselland» wird auf Ende Jahr eingestellt [Internet]. [cited 2025 May 10]. Available from: https://www.baselland.ch/politik-und-behorden/direktionen/volkswirtschafts-und-gesundheitsdirektion/medienmitteilungen/projekt-breites-testen-baselland-wird-auf-ende-jahr-eingestellt

27. rts.ch [infoSport] [Internet]. 2021 [cited 2025 Apr 17]. Le point sur les mesures contre le Covid dans les écoles canton par canton. Available from: https://www.rts.ch/info/regions/12409503-le-point-sur-les-mesures-contre-le-covid-dans-les-ecoles-canton-par-canton.html

28. Littlecotta H, Krishnaratnea S, Burns J, Rehfuess E, Sell K, Klinger C, et al. Measures implemented in the school setting to contain the COVID-19 pandemic. Cochrane Database of Systematic Reviews. 2024;(5). doi:10.1002/14651858.CD015029.pub2

29. Stehlé J, Voirin N, Barrat A, Cattuto C, Isella L, Pinton JF, et al. High-resolution measurements of face-to-face contact patterns in a primary school. PloS one. 2011;6(8):e23176.

30. Banholzer N, Bittel P, Jent P, Furrer L, Zürcher K, Egger M, et al. Molecular detection of SARS-CoV-2 and other respiratory viruses in saliva and classroom air: a two winters tale. Clinical Microbiology and Infection. 2024;30(6):829–e1.

31. Basel Landschaft Kantonaler Kriesenstab. Einverständniserklärung Schule Eltern. 2021.

32. COVID-19 Switzerland Coronavirus Dashboard [Internet]. [cited 2024 Jul 11]. Available from: https://www.covid19.admin.ch/en/overview

33. statistique O fédéral de la. Régions MedStat [Internet]. [cited 2024 Mar 18]. Available from: https://www.bfs.admin.ch/bfs/fr/home/statistiken/gesundheit/nomenklaturen/medsreg.html

34. Federal Statistical Office. Niveaux géographiques - résultats [Internet]. 2024 [cited 2025 Apr 9]. Available from: https://www.agvchapp.bfs.admin.ch/fr/typologies/results?SnapshotDate=01.01.2024&SelectedTypologies%5B0%5D=HR_GDETYP2020

35. Federal Statistical Office. Regional portraits 2021: key data of all communes 2004-2020 [Internet]. 2021 [cited 2025 Apr 9]. Available from: https://www.bfs.admin.ch/asset/en/15864443

36. Coelho FC, Araújo EC, Keiser O. COVID-19 in Switzerland real-time epidemiological analyses powered by EpiGraphHub. Sci Data. 2022 Nov 17;9(1):707. doi:10.1038/s41597-022-01813-5

37. König V. Compulsory education in the Canton of Basel-Landschaft.

38. Calmon L, Colosi E, Bassignana G, Barrat A, Colizza V. Preserving friendships in school contacts: An algorithm to construct synthetic temporal networks for epidemic modelling. PLOS Computational Biology. 2024 Dec 9;20(12):e1012661. doi:10.1371/journal.pcbi.1012661

39. Colosi E, Bassignana G, Contreras DA, Poirier C, Boëlle PY, Cauchemez S, et al. Screening and vaccination against COVID-19 to minimise school closure: a modelling study. The Lancet Infectious Diseases. 2022;22(7):977–89.

40. Colosi E, Bassignana G, Barrat A, Lina B, Vanhems P, Bielicki J, et al. Minimising school disruption under high incidence conditions due to the Omicron variant in France, Switzerland, Italy, in January 2022. Eurosurveillance. 2023 Feb 2;28(5):2200192. doi:10.2807/1560-7917.ES.2023.28.5.2200192

41. Colosi E, Bassignana G, Barrat A, Colizza V. Modelling COVID-19 in school settings to evaluate prevention and control protocols. Anaesthesia Critical Care & Pain Medicine. 2022 Apr 1;41(2):101047. doi:10.1016/j.accpm.2022.101047

42. Colosi E, Lina B, Elias C, Vanhems P, Colizza V. Proactive vs. reactive COVID-19 screening in schools: Lessons from experimental protocols in France during the Delta and Omicron waves. PNAS Nexus. 2026 Mar 9;pgag061. doi:10.1093/pnasnexus/pgag061

43. Young BE, Ong SWX, Kalimuddin S, Low JG, Tan SY, Loh J, et al. Epidemiologic Features and Clinical Course of Patients Infected With SARS-CoV-2 in Singapore. JAMA. 2020 Apr 21;323(15):1488. doi:10.1001/jama.2020.3204

44. He X, Lau EHY, Wu P, Deng X, Wang J, Hao X, et al. Temporal dynamics in viral shedding and transmissibility of COVID-19. Nat Med. 2020 May;26(5):672–5. doi:10.1038/s41591-020-0869-5

45. Hart WS, Miller E, Andrews NJ, Waight P, Maini PK, Funk S, et al. Generation time of the alpha and delta SARS-CoV-2 variants: an epidemiological analysis. The Lancet Infectious Diseases. 2022 May;22(5):603–10. doi:10.1016/S1473-3099(22)00001-9

46. Goldstein E, Lipsitch M, Cevik M. On the Effect of Age on the Transmission of SARS-CoV-2 in Households, Schools, and the Community. The Journal of Infectious Diseases. 2021 Feb 13;223(3):362–9. doi:10.1093/infdis/jiaa691

47. Davies NG, Klepac P, Liu Y, Prem K, Jit M, CMMID COVID-19 working group, et al. Age-dependent effects in the transmission and control of COVID-19 epidemics. Nat Med. 2020 Aug;26(8):1205– 11. doi:10.1038/s41591-020-0962-9

48. Fontanet A, Tondeur L, Grant R, Temmam S, Madec Y, Bigot T, et al. SARS-CoV-2 infection in schools in a northern French city: a retrospective serological cohort study in an area of high transmission, France, January to April 2020. Eurosurveillance. 2021 Apr 15;26(15). doi:10.2807/1560-7917.ES.2021.26.15.2001695

49. Dattner I, Goldberg Y, Katriel G, Yaari R, Gal N, Miron Y, et al. The role of children in the spread of COVID-19: Using household data from Bnei Brak, Israel, to estimate the relative susceptibility and infectivity of children. Lloyd-Smith J, editor. PLoS Comput Biol. 2021 Feb 11;17(2):e1008559. doi:10.1371/journal.pcbi.1008559

50. Han MS, Choi EH, Chang SH, Jin BL, Lee EJ, Kim BN, et al. Clinical Characteristics and Viral RNA Detection in Children With Coronavirus Disease 2019 in the Republic of Korea. JAMA Pediatr. 2021 Jan 1;175(1):73. doi:10.1001/jamapediatrics.2020.3988

51. Smith LE, Potts HWW, Amlôt R, Fear NT, Michie S, Rubin GJ. Adherence to the test, trace, and isolate system in the UK: results from 37 nationally representative surveys. BMJ. 2021 Mar 31;608. doi:10.1136/bmj.n608

52. Powell AA, Kirsebom F, Stowe J, Ramsay ME, Lopez-Bernal J, Andrews N, et al. Protection against symptomatic infection with delta (B.1.617.2) and omicron (B.1.1.529) BA.1 and BA.2 SARS-CoV-2 variants after previous infection and vaccination in adolescents in England, August, 2021–March, 2022: a national, observational, test-negative, case-control study. The Lancet Infectious Diseases. 2023 Apr;23(4):435–44. doi:10.1016/S1473-3099(22)00729-0

53. Stringhini S, Zaballa ME, Pullen N, Perez-Saez J, De Mestral C, Loizeau AJ, et al. Seroprevalence of anti-SARS-CoV-2 antibodies 6 months into the vaccination campaign in Geneva, Switzerland, 1 June to 7 July 2021. Eurosurveillance. 2021 Oct 28;26(43). doi:10.2807/1560-7917.ES.2021.26.43.2100830

54. Tancredi S, Chiolero A, Wagner C, Haller ML, Chocano-Bedoya P, Ortega N, et al. Seroprevalence trends of anti-SARS-CoV-2 antibodies and associated risk factors: a population-based study. Infection. 2023 Oct;51(5):1453–65. doi:10.1007/s15010-023-02011-0

55. Eyre DW, Taylor D, Purver M, Chapman D, Fowler T, Pouwels KB, et al. Effect of Covid-19 Vaccination on Transmission of Alpha and Delta Variants. N Engl J Med. 2022 Feb 24;386(8):744–56. doi:10.1056/NEJMoa2116597

56. Daon Y, Huppert A, Obolski U. An accurate model for SARS-CoV-2 pooled RT-PCR test errors. Royal Society Open Science. 2021 Nov 3;8(11):210704. doi:10.1098/rsos.210704

57. Migueres M, Vellas C, Abravanel F, Da Silva I, Dimeglio C, Ferrer V, et al. Testing individual and pooled saliva samples for sars-cov-2 nucleic acid: a prospective study. Diagnostic Microbiology and Infectious Disease. 2021 Nov 1;101(3):115478. doi:10.1016/j.diagmicrobio.2021.115478

58. Hofman P, Allegra M, Salah M, Benzaquen J, Tanga V, Bordone O, et al. Evaluation of Sample Pooling for SARS-CoV-2 Detection in Nasopharyngeal Swab and Saliva Samples with the Idylla SARS-CoV-2 Test. Microbiology Spectrum. 2021 Nov 10;9(3):e00996–21. doi:10.1128/Spectrum.00996-21

59. Smith RL, Gibson LL, Martinez PP, Ke R, Mirza A, Conte M, et al. Longitudinal Assessment of Diagnostic Test Performance Over the Course of Acute SARS-CoV-2 Infection. The Journal of Infectious Diseases. 2021 Sep 17;224(6):976–82. doi:10.1093/infdis/jiab337

60. McGee RS, Homburger JR, Williams HE, Bergstrom CT, Zhou AY. Model-driven mitigation measures for reopening schools during the COVID-19 pandemic. Proc Natl Acad Sci U S A. 2021 Sep 28;118(39):e2108909118. doi:10.1073/pnas.2108909118 PubMed PMID: 34518375; PubMed Central PMCID: PMC8488607.

61. Torneri A, Willem L, Colizza V, Kremer C, Meuris C, Darcis G, et al. Controlling SARS-CoV-2 in schools using repetitive testing strategies. Leung K, van der Meer JW, Young BC, editors. eLife. 2022 Jul 5;11:e75593. doi:10.7554/eLife.75593

62. Perez-Saez J, Bellon M, Lessler J, Berthelot J, Hodcroft EB, Michielin G, et al. Evolving infectious disease dynamics shape school-based intervention effectiveness. Nat Commun. 2025 Jul 17;16(1):6597. doi:10.1038/s41467-025-61925-5

63. Hemani G, Thomas AC, Walker JG, Trickey A, Nixon E, Ellis D, et al. Modelling pooling strategies for SARS-CoV-2 testing in a university setting. Wellcome Open Res. 2021 Mar 30;6:70. doi:10.12688/wellcomeopenres.16639.1

64. Sewell DK. Leveraging Network Structure to Improve Pooled Testing Efficiency. Journal of the Royal Statistical Society Series C: Applied Statistics. 2022 Nov 1;71(5):1648–62. doi:10.1111/rssc.12594

65. Johnson KE, Pasco R, Woody S, Lachmann M, Johnson-Leon M, Bhavnani D, et al. Optimizing COVID-19 testing strategies on college campuses: Evaluation of the health and economic costs. PLoS Comput Biol. 2023 Dec;19(12):e1011715. doi:10.1371/journal.pcbi.1011715 PubMed PMID: 38134223; PubMed Central PMCID: PMC10773932.

66. Callaghan T, Lueck JA, Trujillo KL, Ferdinand AO. Rural and Urban Differences in COVID-19 Prevention Behaviors. The Journal of Rural Health. 2021;37(2):287–95. doi:10.1111/jrh.12556

67. Dohle S, Wingen T, Schreiber M. Acceptance and Adoption of Protective Measures During the COVID-19 Pandemic: The Role of Trust in Politics and Trust in Science. Social Psychological Bulletin. 2020 Dec 23;15(4):1–23. doi:10.32872/spb.4315

68. Wachira E, Laki K, Chavan B, Aidoo-Frimpong G, Kingori C. Factors Influencing COVID-19 Prevention Behaviors. J of Prevention. 2023 Feb 1;44(1):35–52. doi:10.1007/s10935-022-00719-7

69. Papageorge NW, Zahn MV, Belot M, van den Broek-Altenburg E, Choi S, Jamison JC, et al. Socio-demographic factors associated with self-protecting behavior during the Covid-19 pandemic. J Popul Econ. 2021 Apr 1;34(2):691–738. doi:10.1007/s00148-020-00818-x

70. Banholzer N, Munday JD, Jent P, Bittel P, Dall’Amico L, Furrer L, et al. The relative contribution of close-proximity contacts, shared classroom exposure and indoor air quality to respiratory virus transmission in schools. Nat Commun. 2025 Nov 27;16(1):11678. doi:10.1038/s41467-025-66719-3

